# Machine learning models predict long COVID outcomes based on baseline clinical and immunologic factors

**DOI:** 10.1101/2025.02.12.25322164

**Authors:** Naresh Doni Jayavelu, Hady Samaha, Sonia Tandon Wimalasena, Annmarie Hoch, Jeremy P Gygi, Gisela Gabernet, Al Ozonoff, Shanshan Liu, Carly E. Milliren, Ofer Levy, Lindsey R. Baden, Esther Melamed, Lauren I. R. Ehrlich, Grace A. McComsey, Rafick P. Sekaly, Charles B. Cairns, Elias K. Haddad, Joanna Schaenman, Albert C. Shaw, David A. Hafler, Ruth R. Montgomery, David B. Corry, Farrah Kheradmand, Mark A. Atkinson, Scott C. Brakenridge, Nelson I Agudelo Higuita, Jordan P. Metcalf, Catherine L. Hough, William B. Messer, Bali Pulendran, Kari C. Nadeau, Mark M. Davis, Linda N. Geng, Ana Fernandez Sesma, Viviana Simon, Florian Krammer, Monica Kraft, Chris Bime, Carolyn S. Calfee, David J. Erle, Charles R. Langelier, IMPACC Network, Leying Guan, Holden T. Maecker, Bjoern Peters, Steven H. Kleinstein, Elaine F. Reed, Joann Diray-Arce, Nadine Rouphael, Matthew C. Altman

**Affiliations:** Clinical and Data Coordinating Center (CDCC) Precision Vaccines Program, Boston Children’s Hospital, Boston, MA 02115, USA; Benaroya Research Institute, University of Washington, Seattle, WA 98101, USA; La Jolla Institute for Immunology, La Jolla, CA 92037, USA; Knocean Inc. Toronto, ON M6P 2T3, Canada; Precision Vaccines Program, Boston Children’s Hospital, Harvard Medical School, Boston, MA 02115, USA; Brigham and Women’s Hospital, Harvard Medical School, Boston, MA 02115, USA; Metabolon Inc, Morrisville, NC 27560, USA; Prevention of Organ Failure (PROOF) Centre of Excellence, University of British Columbia, Vancouver, BC V6T 1Z3, Canada; Case Western Reserve University and University Hospitals of Cleveland, Cleveland, OH 44106, USA; Drexel University, Tower Health Hospital, Philadelphia, PA 19104, USA; MyOwnMed Inc., Bethesda, MD 20817, USA; Emory School of Medicine, Atlanta, GA 30322, USA; Icahn School of Medicine at Mount Sinai, New York, NY 10029, USA; Immunai Inc. New York, NY 10016, USA; Oregon Health Sciences University, Portland, OR 97239, USA; Stanford University School of Medicine, Palo Alto, CA 94305, USA; David Geffen School of Medicine at the University of California Los Angeles, Los Angeles CA 90095, USA; University of California San Francisco, San Francisco, CA 94115, USA; Yale School of Medicine, New Haven, CT 06510, USA; Yale School of Public Health, New Haven, CT 06510, USA; Baylor College of Medicinea and the Center for Translational Research on Inflammatory Diseases, Houston, TX 77030, USA; Oklahoma University Health Sciences Center, Oklahoma City, OK 73104, USA; University of Arizona, Tucson AZ 85721, USA; University of Florida, Gainesville, FL 32611, USA; University of Florida, Jacksonville, FL 32218, USA; University of South Florida, Tampa FL 33620, USA; The University of Texas at Austin, Austin, TX 78712, USA

**Keywords:** COVID-19, SARS-CoV-2, PASC, Long COVID, Patient Reported Outcomes, modeling

## Abstract

The post-acute sequelae of SARS-CoV-2 (PASC), also known as long COVID, remain a significant health issue that is incompletely understood. Predicting which acutely infected individuals will go on to develop long COVID is challenging due to the lack of established biomarkers, clear disease mechanisms, or well-defined sub-phenotypes. Machine learning (ML) models offer the potential to address this by leveraging clinical data to enhance diagnostic precision. We utilized clinical data, including antibody titers and viral load measurements collected at the time of hospital admission, to predict the likelihood of acute COVID-19 progressing to long COVID. Our machine learning models achieved median AUROC values ranging from 0.64 to 0.66 and AUPRC values between 0.51 and 0.54, demonstrating their predictive capabilities. Feature importance analysis revealed that low antibody titers and high viral loads at hospital admission were the strongest predictors of long COVID outcomes. Comorbidities, including chronic respiratory, cardiac, and neurologic diseases, as well as female sex, were also identified as significant risk factors for long COVID. Our findings suggest that ML models have the potential to identify patients at risk for developing long COVID based on baseline clinical characteristics. These models can help guide early interventions, improving patient outcomes and mitigating the long-term public health impacts of SARS-CoV-2.

## INTRODUCTION

The definition of post-acute sequelae of SARS-CoV-2 (PASC), or post-COVID-19 syndrome (Long COVID) has been evolving since its initial characterization. The CDC defines long COVID as multiple overlapping physical and cognitive symptoms that present four weeks or more after the onset of infection, and persistent for weeks, months or years[1]. One in seven US adults have experienced at least one symptom of long COVID[2], while the UK estimates 2 million individuals have been affected by this condition[3]. It is challenging to determine which factors are predictive of PASC[4]. Few studies have attempted to use machine learning to create a predictive model of PASC using electronic healthcare records and have differed in study population characteristics and definition of PASC[5,6]. Factors such as age, sex, specific symptoms, and co-morbidities as well as health utilization rates could represent predictive factors. However, published models have been based on non-curated data from healthcare records have not included laboratory data specific to SARS-CoV-2 such as antibody levels and viral loads. Here, we present a predictive model of PASC using a large and well characterized cohort of patients with COVID-19 admitted to hospitals across the US early in the pandemic (the IMmuno Phenotyping Assessment in a COVID-19 Cohort, IMPACC) and integrate demographic, clinical and laboratory data collected at the time of hospitalization for acute infection

## RESULTS

### Demographics and descriptive statistics

1,164 participants were enrolled between May 5^th^, 2020 and March 19^th^, 2021 and followed up to 28 days post hospitalization, then at quarterly intervals for up to 12 months post hospital dischargeAs previously reported, 590/702 (84%) who were alive at least 3 months post hospital discharge completed at least one quarterly set of symptom and patient-reported outcome (PRO) surveys. This group differed from non-responders to the surveys by having shorter hospitalization or fewer limitations at discharge (42% versus 57%). PASC was defined in this cohort as previously described with patient reported outcomes of clinical deficits (Ozonoff et al 2024)[7]. Data from 385 participants was available for modeling after removing missing data. Of these 385, 238 (61.8%) had minimal or no deficit by PRO, while 147 (38.2%) did have a significant deficit and were therefore labeled as long COVID. Table 1 details the demographics, clinical characteristics, baseline radiographic and laboratory findings of this subset of the cohort. Median age was 57 years (IQR 20.0) and did not differ between those with and without evidence of long COVID. The analysis cohort was predominantly male (n=241; 63%), though more females than males reported long COVID (47% vs. 33%, p=0.005). Slightly more participants reporting long COVID were white 59% vs 52%, p=0.049), but ethnicity did not differ significantly. 93% of participants had at least one comorbidity, and the presence of a co-morbidity was higher in those with long COVID (p<0.001). Hypertension, diabetes mellitus, chronic lung disease (not asthma) and chronic cardiac disease were the most common comorbid conditions reported, and all were more prevalent in participants classified as having long COVID. The majority of the analysis cohort (86%; n=332) had a body mass index (BMI) above 25 kg/m^2,^, and class 3 obesity (BMI>40) was more commonly seen in the participants with long COVID (20% vs 11%, p=0.041). Upon admission, 157 (41%) had an elevated baseline C-reactive protein (CRP) (≥10 mg/L) and 280 (73%) had infiltrate(s) on chest imaging, but there were no differences between those with long COVID and those without. Similarly, the majority required some ventilatory or oxygenation support on admission (79% or n=305) but baseline gas exchange and respiratory ordinal score were not associated with evidence of long COVID. Use of steroids and, remdesivir also were not associated with long COVID prevalence, nor was the prevalence of complications during the hospital stays. 186 (48%) out of 385 reported at least one symptom during the quarterly surveys, most commonly cardiopulmonary (cough or dyspnea) (33%), followed by gastrointestinal, neurologic or upper respiratory (32%) or systemic (fever, fatigue, myalgia, chills) (26%), and all reported symptoms were more common in those with long COVID as defined by PROs. As the study was conducted between May 2020 and March 2021, none of the participants had received a COVID-19 vaccine prior to admission.

**Table 1:** Demographics, Clinical Characteristics, Comorbidities, Radiographic Findings and Laboratory Testing of Cohort Participants at Baseline (N=385)

|  |  | Overall<br>(n=385) | Minimal<br>Cluster<br>(n=238) | Deficit<br>Cluster<br>(n=147) | Overall p-<br>value |
| --- | --- | --- | --- | --- | --- |
| Age at enrollment (years), median (IQR) (n=385) |  | 57.0<br>(20.0) | 56.0 (21.0) | 57.0 (18.0) | 0.107 |
| Sex at birth | Male | 241<br>(63%) | 162 (68%) | 79 (54%) | 0.005 |
|  | Female | 144<br>(37%) | 76 (32%) | 68 (46%) |  |
| Race | White | 210<br>(55%) | 124 (52%) | 86 (59%) | 0.049 |
|  | Black/African<br>American | 70<br>(18%) | 42 (18%) | 28 (19%) |  |
|  | Asian | 15 (4%) | 13 (5%) | 2 (1%) |  |
| Ethnicity | Non-Hispanic | 254<br>(66%) | 156 (66%) | 98 (67%) | 0.634 |
|  | Hispanic | 119<br>(31%) | 73 (31%) | 46 (31%) |  |
| Comorbidities | None | 27 (7%) | 23 (10%) | 4 (3%) | <.001 |
|  | Hypertension | 206<br>(54%) | 116 (49%) | 90 (61%) | 0.017 |
|  | Diabetes | 131<br>(34%) | 69 (29%) | 62 (42%) | 0.008 |
|  | Chronic lung disease | 62<br>(16%) | 28 (12%) | 34 (23%) | 0.003 |
|  | Asthma | 59<br>(15%) | 34 (14%) | 25 (17%) | 0.471 |
|  | Chronic cardiac<br>disease | 89<br>(23%) | 42 (18%) | 47 (32%) | 0.001 |
|  | Chronic kidney disease | 38<br>(10%) | 24 (10%) | 14 (10%) | 0.858 |
|  | Liver disease | 21 (5%) | 12 (5%) | 9 (6%) | 0.65 |
|  | Chronic neurological<br>disorder | 42<br>(11%) | 12 (5%) | 30 (20%) | <.001 |
|  | History of SOT or BMT | 24 (6%) | 12 (5%) | 12 (8%) | 0.218 |
|  | HIV or AIDS | 4 (1%) | 2 (1%) | 2 (1%) | 0.625 |
|  | Malignant neoplasm | 32 (8%) | 22 (9%) | 10 (7%) | 0.399 |
|  | Substance use (drugs,<br>alcohol, and/or<br>cannabis) | 22 (6%) | 12 (5%) | 10 (7%) | 0.47 |
|  | Current or former | 116 | 69 (29%) | 47 (32%) | 0.536 |
|  | smoking and/or vaping | (30%) |  |  |  |
| BMI Category | Overweight | 109<br>(28%) | 74 (31%) | 35 (24%) | 0.041 |
|  | Class 1-2 Obesity (30-39.9) | 169<br>(44%) | 102 (43%) | 67 (46%) |  |
|  | Class 3 Obesity (40+) | 54<br>(14%) | 25 (11%) | 29 (20%) |  |
| Symptom onset to hospitalization (days), median (IQR) (n=330) |  | 7.0 (6.0) | 7.0 (6.0) | 7.0 (7.0) | 0.349 |
| Symptom onset to hospitalization | ≤3 | 81<br>(21%) | 50 (21%) | 31 (21%) | 0.489 |
|  | 4-7 | 101<br>(26%) | 59 (25%) | 42 (29%) |  |
|  | 8-14 | 125<br>(32%) | 83 (35%) | 42 (29%) |  |
|  | >14 | 23 (6%) | 16 (7%) | 7 (5%) |  |
| Length of stay (days), median (IQR) (n=365) |  | 5.0 (5.0) | 5.0 (5.0) | 5.0 (5.0) | 0.914 |
| Level of respiratory support | Mechanically ventilated or ECMO | 31 (8%) | 19 (8%) | 12 (8%) | 0.084 |
|  | Non-invasive ventilation or high flow | 61<br>(16%) | 44 (18%) | 17 (12%) |  |
|  | Supplemental oxygen (not high flow) | 213<br>(55%) | 134 (56%) | 79 (54%) |  |
|  | None | 80<br>(21%) | 41 (17%) | 39 (27%) |  |
| SpO2/FiO2 ratio category at lowest sat | 235 or lower | 69<br>(18%) | 50 (21%) | 19 (13%) | 0.253 |
|  | 236-315 | 71<br>(18%) | 43 (18%) | 28 (19%) |  |
|  | 315 or higher | 223<br>(58%) | 132 (55%) | 91 (62%) |  |
| SOFA Score, median (IQR) |  | 0.0 (2.0) | 0.0 (2.0) | 0.0 (2.0) | 0.381 |
| Infiltrates on CXR or CT | No infiltrates | 85<br>(23%) | 50 (22%) | 35 (25%) | 0.568 |
|  | Unilateral infiltrates | 31 (8%) | 21 (9%) | 10 (7%) |  |
|  | Bilateral infiltrates | 249<br>(68%) | 153 (68%) | 96 (68%) |  |
| Abnormal Lymphocyte count | (<500/microliter) | 48<br>(12%) | 28 (12%) | 20 (14%) | 0.595 |
| Abnormal Platelets | (<100,000/microliter) | 14 (4%) | 8 (3%) | 6 (4%) | 0.714 |
| Abnormal D-dimer | (>0.5 mg/L) | 192<br>(50%) | 126 (53%) | 66 (45%) | 0.125 |
| Abnormal ALT | (>1.5x site-specific | 67 | 48 (20%) | 19 (13%) | 0.069 |
|  | ULN) | (17%) |  |  |  |
| Abnormal Creatinine | (≥1.5 mg/dL) | 46<br>(12%) | 28 (12%) | 18 (12%) | 0.888 |
| Abnormal CRP | (≥10 mg/L) | 157<br>(41%) | 96 (40%) | 61 (41%) | 0.822 |
| Abnormal Troponin | (>0.4 ng/mL) | 29 (8%) | 18 (8%) | 11 (7%) | 0.977 |
| Any complications | Yes | 329<br>(85%) | 209 (88%) | 120 (82%) | 0.095 |
| Remdesivir use | Yes | 251<br>(65%) | 162 (68%) | 89 (61%) | 0.132 |
| Steroids use | Yes | 262<br>(68%) | 168 (71%) | 94 (64%) | 0.174 |
| Convalescent symptoms | Any cardiopulmonary (cough, dyspnea) | 126<br>(33%) | 62 (26%) | 64 (44%) | <.001 |
|  | Any systemic (fever, fatigue, myalgia, chills) | 100<br>(26%) | 38 (16%) | 62 (43%) | <.001 |
|  | Any gastrointestinal, neurologic, or upper respiratory | 121<br>(32%) | 54 (23%) | 67 (47%) | <.001 |
|  | Number of symptom categories present, median (IQR)(n=381) | 0.0 (2.0) | 0.0 (1.0) | 1.0 (2.0) | <.001 |

<sup>1</sup>23% other/declined/unknown/missing; <sup>2</sup>less than 5% missing data; <sup>3</sup>14% missing data; <sup>4</sup>less than 10%
missing data, <sup>5</sup>not including asthma
p-value from chi-square test for categorical variables and Kruskal-Wallis test for continuous variables (age, SOFA score).

### Machine learning models using variables collected at the time of COVID-19 hospitalization predict risk of developing long COVID

We developed a series of machine learning models for predicting the risk of developing a long COVID phenotype versus not developing a long COVID phenotype[7]. Models were built based on the clinical variables, symptoms, antibody (anti-Spike IgG) and viral load (N1 Ct) measurements collected at the time of hospital admission (baseline) for acute SARS-CoV-2 infection. A schematic of the machine learning model development pipeline is shown in Figure 1. First, data were split into stratified training and testing datasets using a random 70:30 split, respectively, where each dataset had equal representation of those with and without subsequent development of a long COVID phenotypes. Next, we selected relevant features by performing feature selection on the training data set using LASSO regression. Models were built on the training data set and model performances were then assessed on the training data and subsequently on the test data set repeated 15 times which was entirely independent from the training data on which the models were constructed. Model performances showed accurate prediction on the training data, as expected, with mean AUROC values ranging from 0.766 to 0.846 (bGLM = 0.780, NNET = 0.766, RF = 0.846, SVM = 0.796, XGB = 0.836) and mean AUPRC (bGLM = 0.688, NNET = 0.669, RF = 0.802, SVM = 0.729, XGB = 0.773) values ranging from 0.669 to 0.802 (supplementary Figure 1). When the models were then tested on the independent test dataset, all five models still achieved a significant predictive performance in terms of mean AUROC, with values ranging from 0.64 to 0.66 (bGLM = 0.657, NNET = 0.658, RF = 0.657, SVM = 0.651, XGB = 0.637). Similarly, AUPRC values ranged from 0.51 to 0.54 (bGLM = 0.526, NNET = 0.531, RF = 0.538, SVM = 0.533, XGB = 0.51) (Figure 2 and supplementary Figure 2). These results demonstrate that a parsimonious set of baseline clinical features can robustly predict subsequent development of a long COVID phenotype.

**Figure 1:**
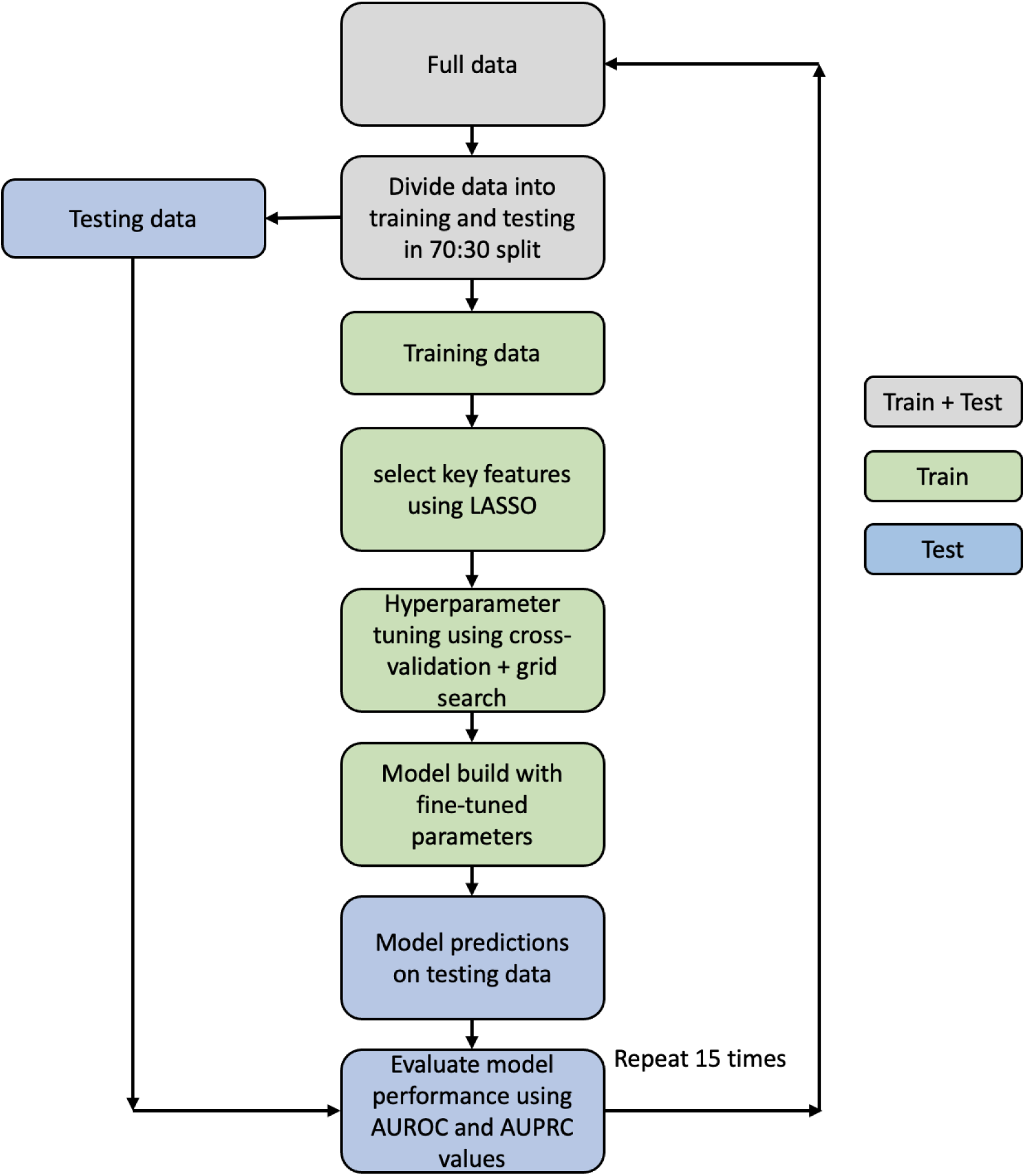
Schematic of the machine learning models development for predicting long COVID.

**Figure 2:**
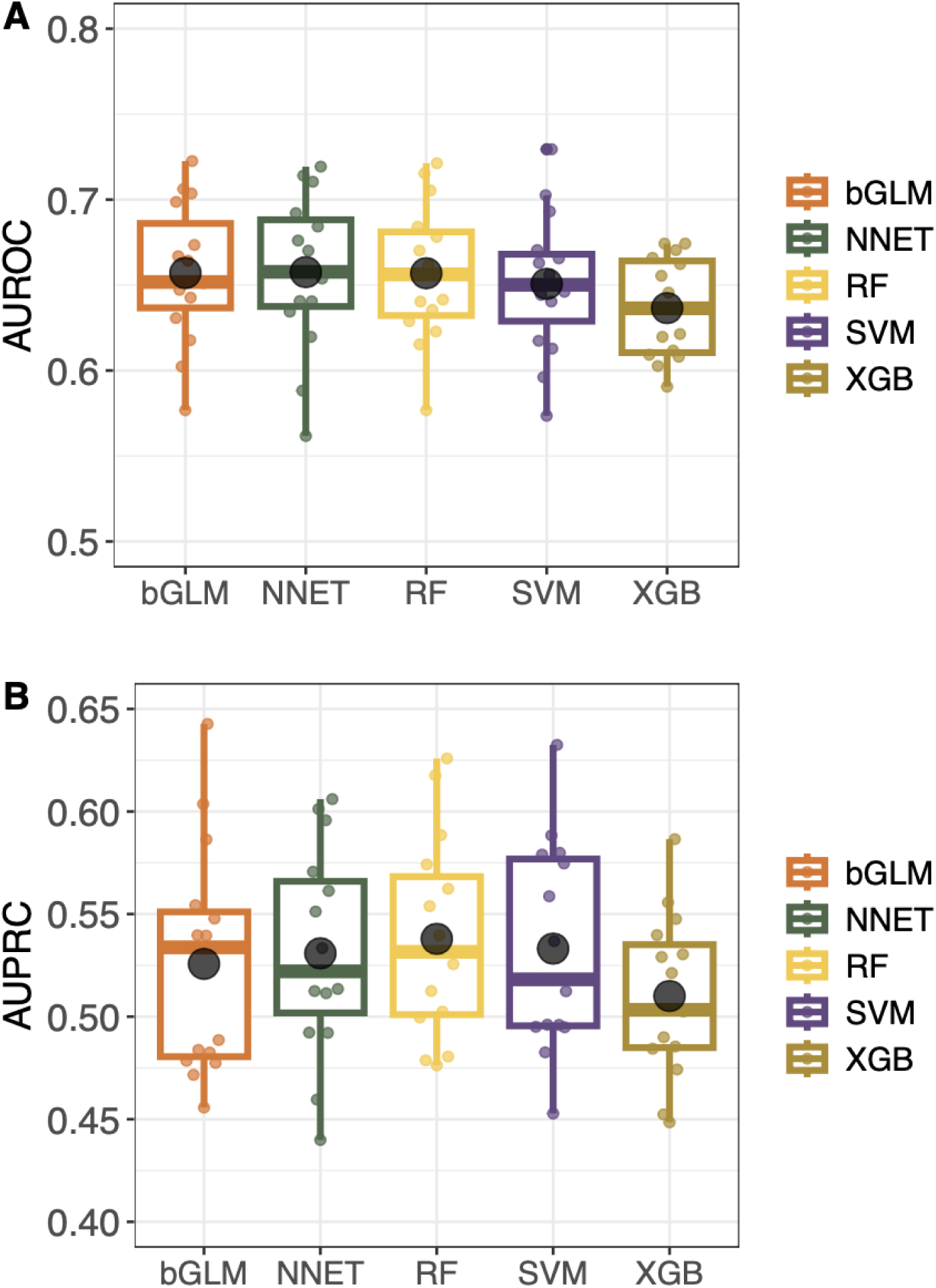
Evaluation of machine learning model’s predictive performance on independent test data for identifying patients at risk of developing a long COVID phenotype. Distribution of (A) AUROC and (B) AUPRC values for all the models.

### Feature importance

The relative importance score of all features for all models were investigated to understand which variables were important to predict long COVID (Figure 3). All models demonstrated that lower antibody titers (anti-Spike IgG) and higher viral loads (N1 Ct) measured at hospital admission were the most statistically important baseline features in identifying patients that go on to develop long COVID symptoms. Among other features, diabetes mellitus, chronic respiratory disease, chronic cardiac disease, chronic neurologic disease and being female were significant risk factors for long COVID. Conversely, symptoms of cough, chest pain, sore throat were indicative of lower risk of developing long COVID.

**Figure 3:**
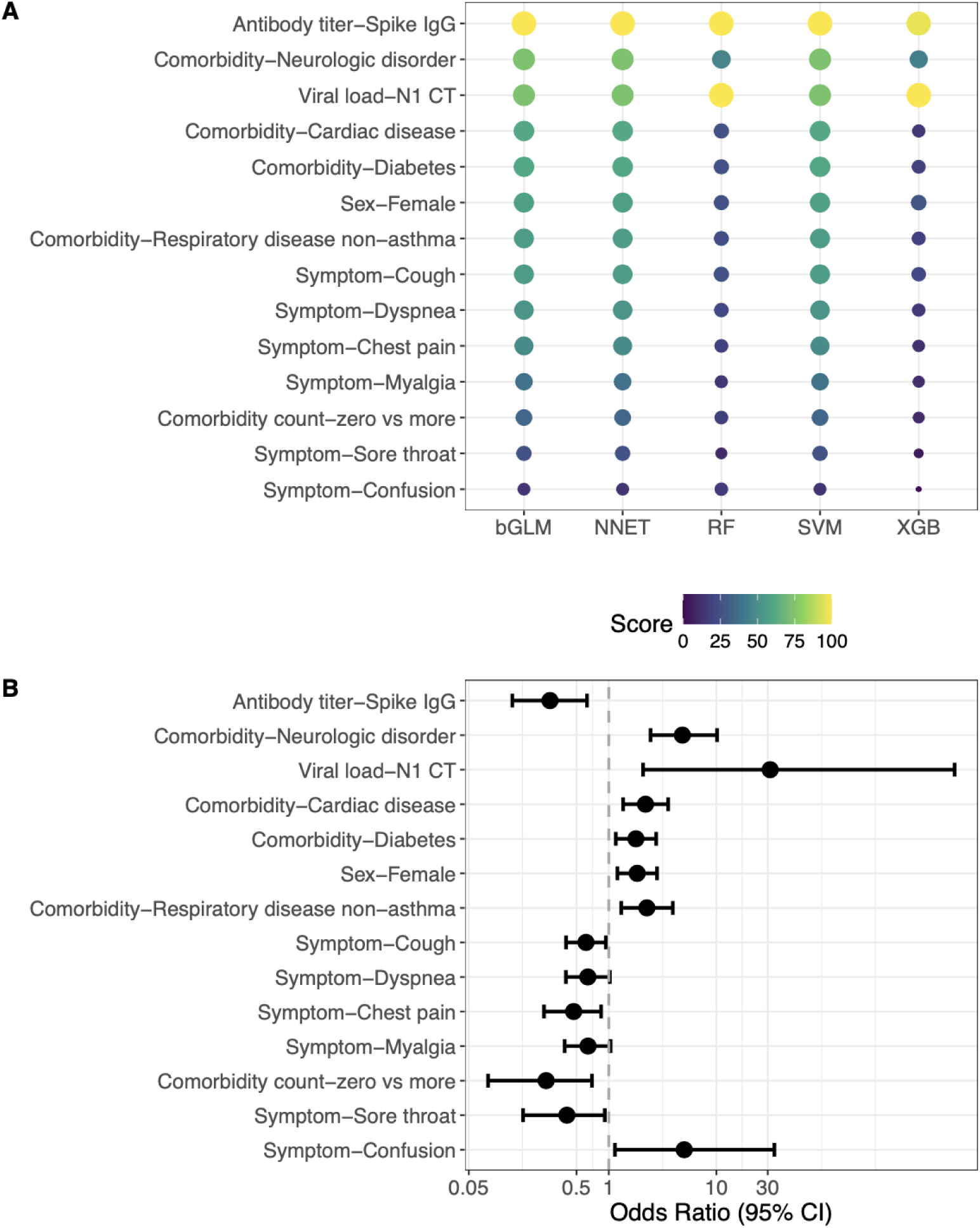
Relative importance of features included in the machine learning models predicting a long COVID phenotype. (A) Dot plot showing scaled importance of all features included in the models predictive of long COVID. The size of the circle shows the relative importance of features. (B) Forest plot showing the univariate model odds ratios for the same features.

## DISCUSSION

Developing a reliable predictive model of long COVID/PASC has proved challenging, largely due to the absence of a specific symptom-based definition for this condition[7–10], unclear understanding of pathophysiology, and absence of validated biomarkers. Four years after the emergence of SARS-CoV-2, few studies have attempted to create a predictive model of long COVID. Antony et al. suggested that factors such as advanced age, female sex, certain comorbidities and medications administered, have a predictive value in determining the likelihood of developing long COVID[5]. This is consistent with the findings of Pfaff et al., who demonstrated that the most important features predictive of long COVID include post-COVID-19 respiratory symptoms, healthcare utilization rates, age as well as selected pre-existing conditions[6]. Finally, Sudre et al. relied on symptoms collected from patients during the early phase of their COVID-19 to determine their likelihood of developing long COVID. They reported that a symptom burden of five or more symptoms during the first week of infection was a strong predictor of PASC. Fatigue, headache, dyspnea, hoarse voice, and myalgia carried the most weight in predicting progression to PASC[4,6]. These studies highlight important components of predicting PASC but are limited in unique ways. The study by Antony et al. lacked diversity and relied on billing codes (likely underestimating PASC to a 0.3% prevalence). The study by Pfaff et al. included patients seeking care in long COVID clinics mostly within the pulmonary department and therefore is not generalizable to different PASC phenotypes. The third study by Sudre et al. was based on self-reported diagnosis and data entered by patients who were app users and limited to spring of 2020. None of these studies integrated clinical laboratory data specific to SARS-CoV-2 such as antibody titer and viral load.

Our study is based on a well-characterized prospective multicenter cohort that includes laboratory values, curated clinical data, as well as patient-reported symptoms and outcomes. Long COVID in the IMPACC cohort previously identified 4 clusters of patients based on reported clinical deficits. The predictive model proposed in this study focuses on baseline SARS-CoV-2 viral load and antibody measurements as features with the highest importance in terms of predictive value of progression to PASC. Many studies have postulated that an elevated viral load at time of COVID-19 positivity is correlated with worse clinical outcomes[11–13] and emerging data suggest an association between viral load and Long COVID[7,14,15]. Moreover, the relationship between viral load and development of complications has already been established for other viruses, especially HIV. An elevated initial viral load on diagnosis of HIV was an accurate predictor of an earlier progression to AIDS[16]. Additionally, a high CSF HIV viral load was one of the most important predictors of progression to dementia in the pre-ART era[16,17]. Nonetheless, according to our data, antiviral therapy (limited to remdesivir in our study) was not associated with a decrease in PASC risk. Passive and active immunization was rarely received in our population (8.3% received convalescent plasma and vaccines were not commonly available), not allowing us to draw any conclusion on the benefit of early antibody in preventing PASC. In addition, steroid use in the acute phase was not associated with a reduction in PASC, and abnormal CRP levels on admission did not differ among those who developed PASC and those who did not.

While certain comorbidities are associated with PASC (e.g., chronic pulmonary, cardiac or neurologic diseases), it is unclear based on our data if some of the PASC disease burden could be misattributed to COVID-19 or if COVID-19 accentuates these pre-existing conditions. We also found that female sex was associated with PASC consistent with other studies[18]. Severity of disease on admission was not associated with PASC in our model, and as noted by others PASC also occurs after mild to moderate COVID[19,20].

Our study has many strengths including the use of advanced ML models to integrate baseline clinical characteristics, antibody titers, and viral loads to predict long COVID outcomes. It enables early identification of individuals at risk, which could allow for more timely interventions and personalized care strategies. The comprehensive feature importance analysis reveals the key predictors, and our data demonstrate robustness of models across multi-center data. We also note the following limitations. First, our cohort was enrolled early in the pandemic and does not fully reflect how COVID-19 currently presents (in patients with hybrid immunity, exposed to variants of concern and having access to more antivirals, introduction of vaccines); in addition, because our study was designed at the onset of the pandemic, we did not comprehensively capture all symptoms that have been assessed to define PASC, though the PRO measures we chose are a reflection of current health status across functional domains and also include a comparison to pre-illness baseline. Second, we did not include additional cohorts (e.g: non-hospitalized COVID-19 cohort nor a hospitalized cohort without COVID-19) and the exclusion of mild to moderate COVID-19 affects the generalizability of our findings. Third, we did not include in our modeling changes of clinical and laboratory data in the acute phase of illness, as our goal was to create a framework for stratification upon presentation to care. Limitation of ML models in predicting long COVID outcomes is their dependance on the quality and comprehensiveness of the input data, which can lead to biased or incomplete predictions if key variables or new symptoms are not captured. Moreover, ML models may struggle with generalizability, as models trained on data from early in the pandemic may not perform as well on data reflecting current patient characteristics, including those with hybrid immunity or exposed to newer variants.

Long COVID has a substantial public health impact. Our study integrates multiple data available at presentation to predict risk of PASC. Our findings suggest that a functional antiviral Ab immune response contributes to viral clearance and may decrease the occurrence of PASC. Our results highlight the benefit of measuring both antibody responses and viral load during the acute phase for the early identification of patients at high risk for PASC, which may facilitate early interventions.

## METHODS

### Ethics

NIAID staff conferred with the Department of Health and Human Services Office for Human Research Protections (OHRP) regarding potential applicability of the public health surveillance exception [45CFR46.102(l) (2)] to the IMPACC study protocol. OHRP concurred that the study satisfied criteria for the public health surveillance exception, and the IMPACC study team sent the study protocol, and participant information sheet for review, and assessment to institutional review boards (IRBs) at participating institutions. Twelve institutions elected to conduct the study as public health surveillance, while 3 sites with prior IRB-approved biobanking protocols elected to integrate and conduct IMPACC under their institutional protocols (The University of Texas at Austin, IRB 2020-04-0117; University of California San Francisco, IRB 20-30497; Case Western Reserve University, IRB STUDY20200573) with informed consent requirements. Participants enrolled under the public health surveillance exclusion were provided information sheets describing the study, samples to be collected, and plans for data de-identification, and use. Those that requested not to participate after reviewing the information sheet were not enrolled.

### Study Design and Setting

The study followed the Strengthening the Reporting of Observational Studies in Epidemiology (STROBE) guidelines for reporting observational studies[21]. The design of the IMPACC study has been previously published[22]. This study was registered at clinicaltrials.gov (NCT0438777). Biologic samples collected consisted of blood and mid-turbinate nasal swabs (self, or staff collected; with step-by-step instructions provided to both staff and study participants). The timepoints were as follows: enrollment (Day 1), and Days 4, 7, 14, 21, and 28 post hospital admission (and if feasible, for discharged participants, Days 14, and 28) and 3, 6, 9 and 12 months after discharge. Only baseline data was used for this modeling. Selected information was available after discharge for self-reported vaccination status, recurrent SARS-CoV-2 infection, and persistence of symptoms, as well as mortality.

### Study Participants

Patients 18 years and older admitted to 20 US hospitals (affiliated with 15 academic institutions) between May 2020 and March 2021 were enrolled within 72 hours of hospital admission for COVID-19 infection. Only confirmed positive SARS-CoV-2 PCR and symptomatic cases attributable to COVID-19 infection were followed longitudinally. Participants did not receive compensation for study participation while inpatient, and subsequently were offered compensation during outpatient follow-up visits. The study was designed to enroll participants of both sexes, and sex at birth was collected based on self-report or care giver report.

### Data Collection and Study Variables

Specific data elements were acquired via review of electronic medical records during the inpatient period: participant demographic characteristics (age, sex at birth, race, ethnicity) comorbidities and body mass index (BMI), presenting signs/symptoms and onset, baseline diagnostic investigations (predefined laboratory values, and radiographic findings), baseline oxygen- and ventilatory-support requirement [either a) not requiring supplemental oxygen, b) requiring oxygen, c) non-invasive ventilation, or high-flow oxygen, or d) invasive mechanical ventilation, and/or extracorporeal membrane oxygenation (ECMO)], relevant clinical outcomes such as length of stay and complications as well as medications used to treat COVID-19 while inpatient.

In addition, self-reported symptoms and standardized patient reported outcome surveys were assessed quarterly for the duration of the study up to 12 months after initial hospital discharge. Patient-reported data was collected using a comprehensive digital remote monitoring tool, in the form of a mobile application developed by *My Own Med, Inc*. Along with the mobile application, an administrative portal was developed to collect information by study personnel during site visits or via telephone interviews by a study coordinator to ensure real-time electronic data capture. The surveys administered at these remote visits included reporting of the following symptoms: upper respiratory symptoms (sore throat, conjunctivitis/red eyes), cardiopulmonary symptoms (shortness of breath (dyspnea), cough), systemic symptoms (fever, chills, fatigue/malaise, muscle aches (myalgia)), neurologic symptoms (loss of smell/taste (anosmia/ageusia), headache), and gastrointestinal symptoms (nausea/vomiting).

In addition, the functional assessments of general health and the evaluation of deficits in specific health domains were conducted using validated Patient-Reported Outcome (PRO) measures, including

⍰ EQ-5D-5L, a standardized, self-administered instrument that describes and quantifies health-related quality of life[23][24–26]
⍰ Patient-Reported Outcomes Measurement Information System (PROMIS). The PROMIS measures administered included:

- *PROMIS® Item Bank v2.0 - Physical Function and PROMIS Item Bank v2.0 -Cognitive Function,* two computer adaptive surveys with tailored questionnaires based on item response theory[27].
- *PROMIS Scale v1.2 - Global Health Mental 2a and* PROMIS Item Bank v1.0 - *Psychosocial Illness Impact – Positive* - Short Form 8a, two surveys with fixed questions[24–26,28]
- *PROMIS Pool v1.0 - Dyspnea Time Extension* computer adaptive instrument for participants who reported shortness of breath[29–31]. This 7-item questionnaire assesses whether there has been a meaningful increase or decrease in the duration of time needed by an adult to perform a given task in the past 7 days compared to 3 months ago due to shortness of breath.

For all PROMIS measures, scoring was based on PROMIS standardized instructions and conversion to a t-statistic[32].

⍰ Health Recovery Score: Overall health was also assessed by a health recovery score utilizing a Visual Analog Scale of 1-100 to indicate overall physical and mental function compared to pre-COVID function.

All data were reviewed centrally to ensure accuracy and consistency. Any data concerns were resolved by querying the site. The full study data collection forms for the quarterly outpatient surveys are provided in the online supplement (Surveys Administered).

Data lock on the survey data was performed on April 7, 2022 and clinical data lock was performed on January 1, 2023.

The full study data collection forms are provided in the online supplement and deidentified data is available upon request.

### Assays

#### SARS-CoV-2 viral load

SARS-CoV-2 viral load was assessed by a central laboratory from nasal swab samples at each time point by RT-PCR of the viral N1, and N2 genes (see online supplement)[33].

#### Anti-SARS-CoV-2 spike (S), and receptor binding domain (RBD) antibodies

Anti-SARS-CoV-2 spike (S), and receptor binding domain (RBD) antibodies were quantified by enzyme-linked immunosorbent assay (ELISA) in serum specimens[34]. (see online supplement)

### Statistics

#### Convalescent clinical outcome assessment

##### Statistical Analysis – Demographic & Clinical variables

We report median (interquartile range, IQR) for continuous variables and frequency (percent) for categorical variables. We examined bivariate associations between demographic and clinical factors and the PRO clusters using Wilcoxon rank sum test for continuous variables and chi-square test for categorical variables. P<0.05 was considered statistically significant.

###### Machine learning modeling

### Feature selection

We used all the features available at the time of hospital admission, including patient demographics, comorbidities, symptoms, baseline clinical characteristics, baseline lab measurements. In addition, we used viral levels (N1 Ct) and antibody measurements (anti-Spike IgG) both collected at the time of (Visit 1) of hospital admission. In total we had access to 93 features. Categorical variables were converted to one-hot encoded. We excluded participants with missing data for any of the variables to ensure the integrity and completeness of the dataset. After this exclusion process, a total of 385 participants remained available for inclusion in the modeling analysis. Feature selection is a commonly used technique in predictive machine learning models to reduce the feature space by removing irrelevant and redundant features. We performed feature selection on only a training data set using LASSO (Least Absolute Shrinkage and Selection Operator) logistic regression. LASSO includes the *l*_1_ norm of feature coefficients as a penalty term to the loss function which forces the coefficients for those features with weak associations to zero. We chose features with non-zero coefficients using 10-fold cross validation. To further reduce the features, we repeated this process 100 times and retained only those features that appeared more than 80 times out of 100 repetitions. We used only those features as input to train all the machine learning models. The features anti-Spike IgG, viral load N1 CT are modeled as continuous variables and all other features as binary variables.

### Outcome variable

We used previously defined Long COVID phenotypes, the four PRO clusters namely MIN, COG, PHY and MLT. We merged the three deficit PRO clusters (COG, PHY and MLT) to represent a long COVD phenotype and the MIN cluster to represent a non-long COVID phenotype. We used this binary variable as a model outcome.

### Machine Learning model development

We trained machine learning models to predict the risk of developing a long COVID phenotype versus non-long COVID phenotype. The R package, *Caret, was* used to train, fine-tune and build final models. Hyperparameters for each model were tuned using *tuneGrid* functionality from caret package with three-fold cross-validation, set to optimize the area under the receiver operating characteristic curve (AUROC). Each model’s performance was evaluated using AUROC and AUPRC (area under precision-recall curve) on set-aside testing data, repeated fifteen times. To interpret the models, we computed the relative importance of all the features included in the model training using *varImp* functionality from caret.

## Data Availability

All data produced are available online at https://www.immport.org/shared/study/SDY1760

## SUPPLEMENTARY FIGURES

**Supplementary Figure 1:**
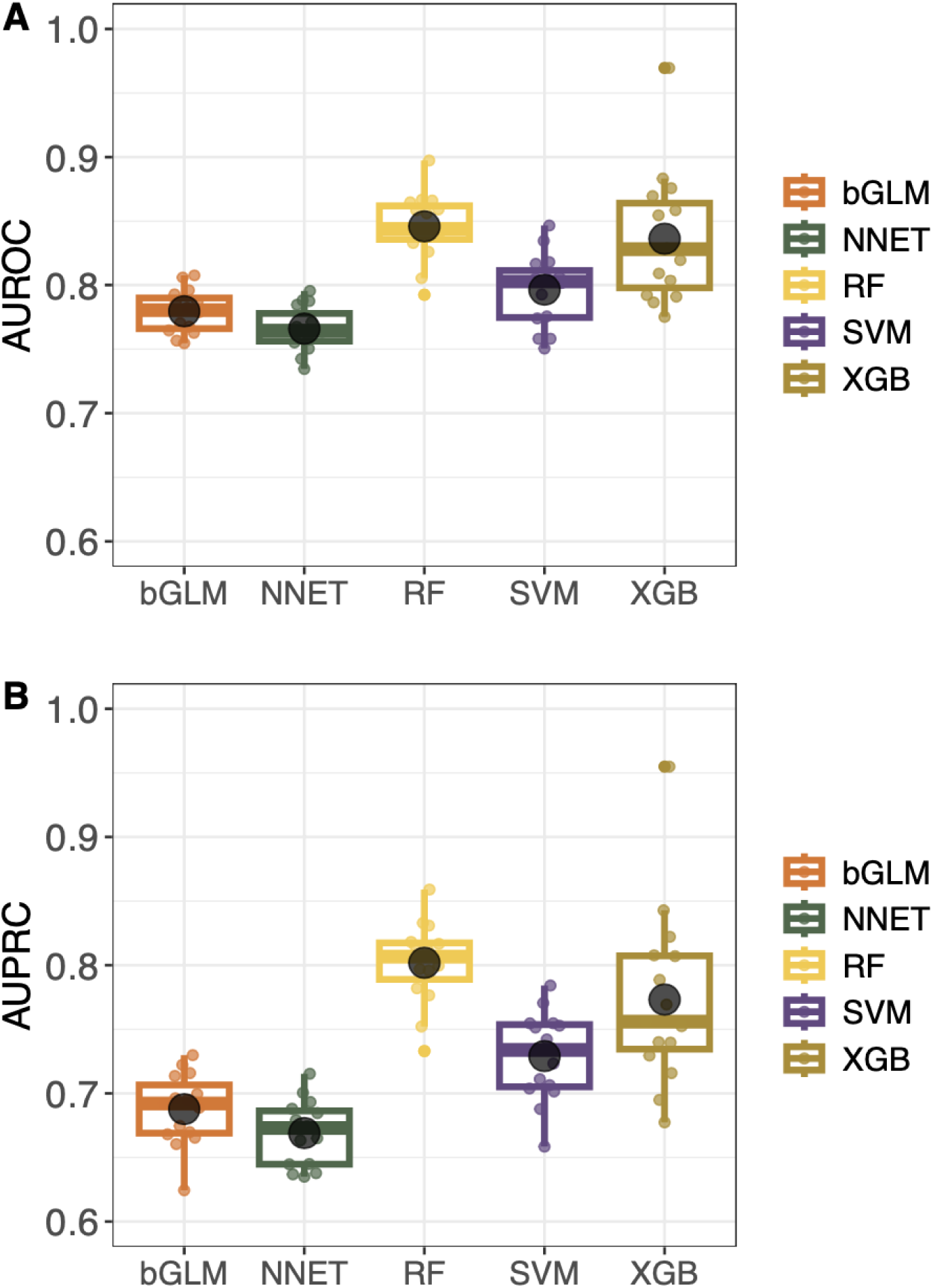
Evaluation of machine learning models predictive performance in identifying patients potentially developing a long COVID phenotype. Distribution of (A) AUROC and (B) AUPRC values across cross validations for all the models.

**Supplementary Figure 2:**
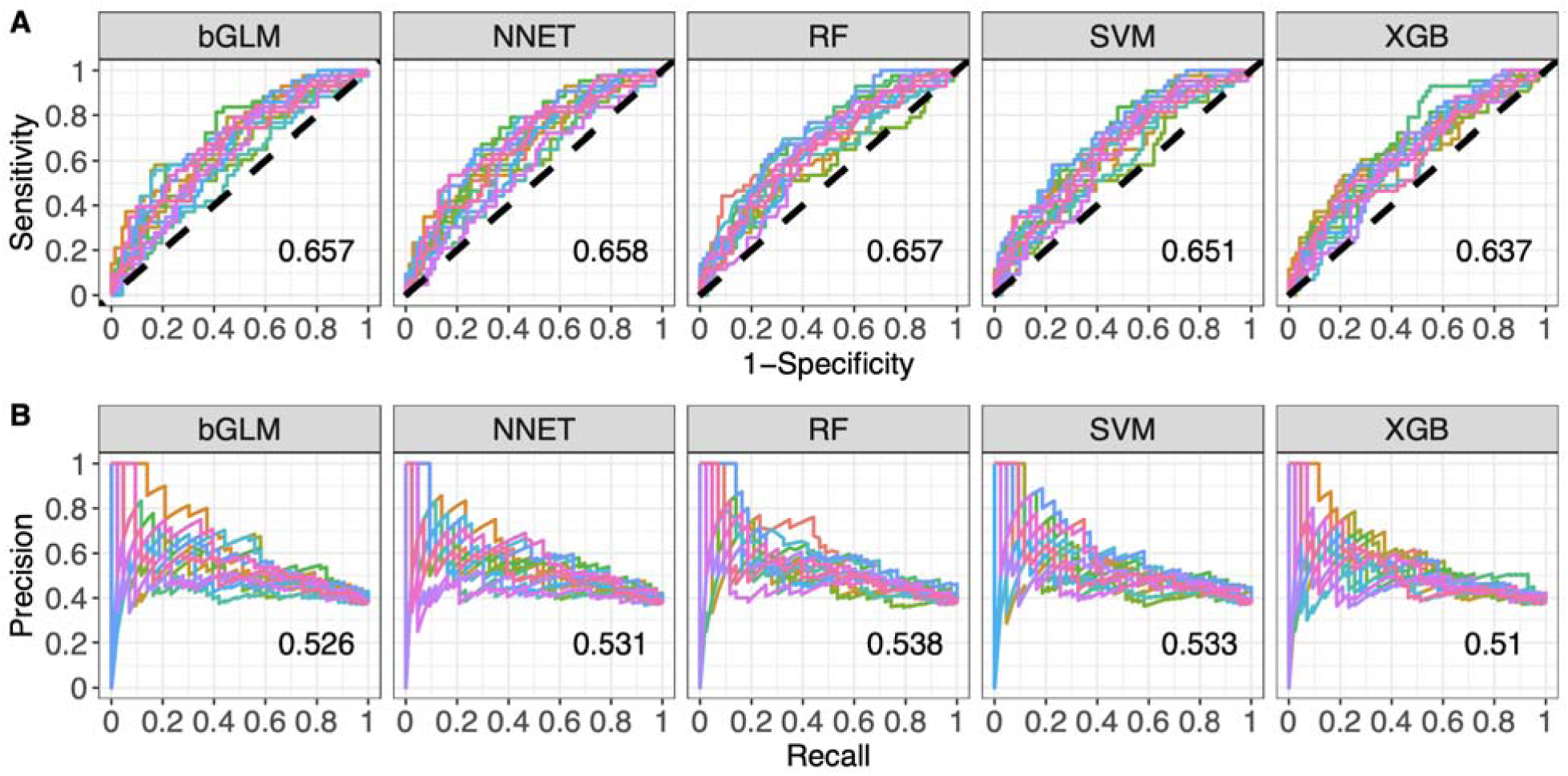
Evaluation of machine learning models predictive performance on test data in identifying patients at the risk of developing a long COVID phenotype. Colored lines denote (A) receiver operating characteristic curves (ROC) and (B) precision-recall curves (PRC) for each test fold with fifteen repeats, evaluating with varying thresholds each model’s predictive ability. The black dotted line indicates the expected ROC curve of a random model.

**Supplementary Figure 3:**
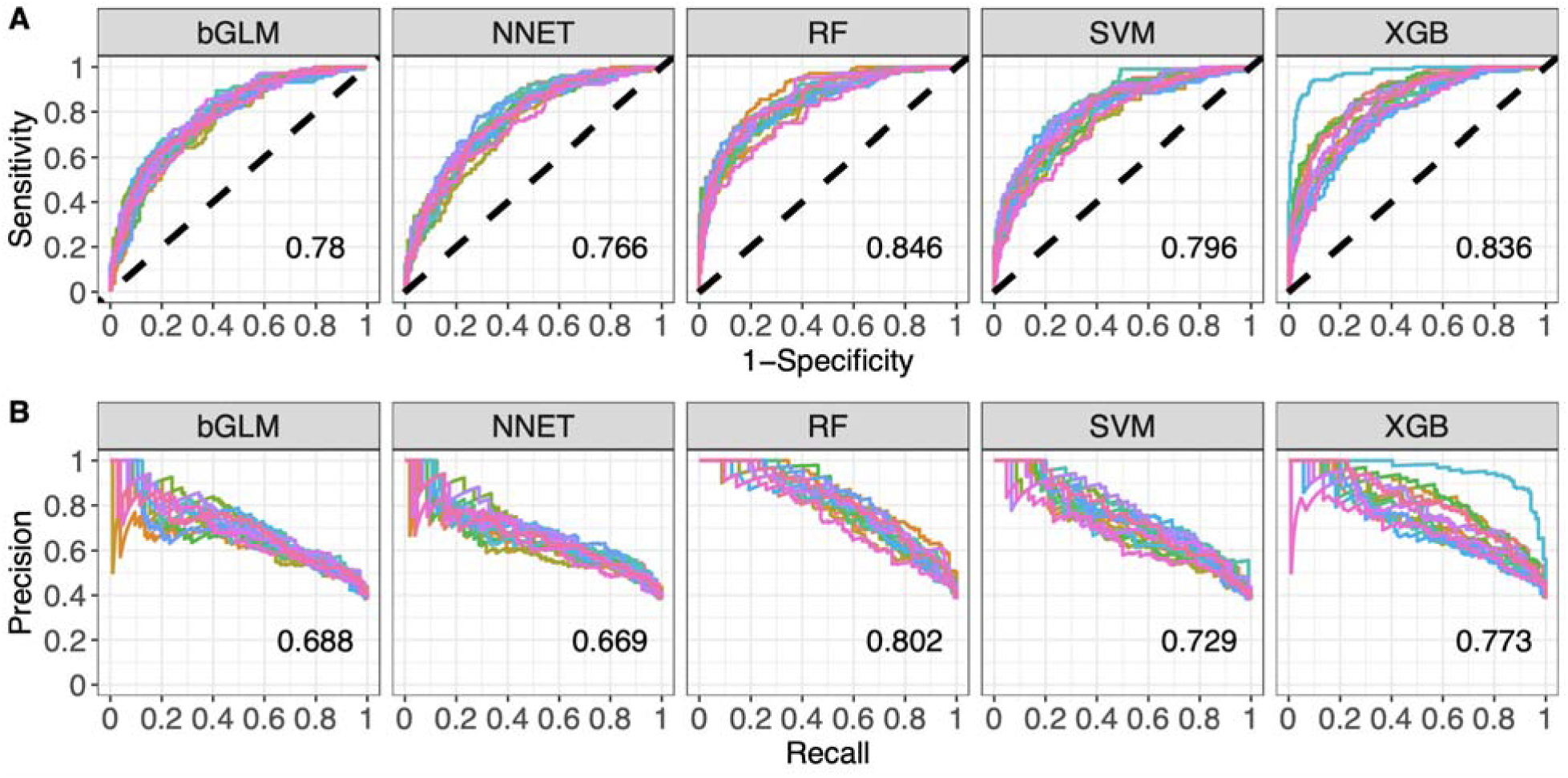
Evaluation of machine learning models predictive performance on train data in identifying patients at the risk of developing a long COVID phenotype. Colored lines denote (A) receiver operating characteristic curves (ROC) and (B) precision-recall curves (PRC) for each train fold with fifteen repeats, evaluating with varying thresholds each model’s predictive ability. The black dotted line indicates the expected ROC curve of a random model.

## DATA AVAILABILITY

The IMPACC Data Sharing Plan is designed to enable the widest dissemination of data, while also protecting the privacy of the participants and the utility of the data by de-identifying and masking potentially sensitive data elements. All IMPACC data including those generated in this study have been deposited in the Immunology Database and Analysis Portal (ImmPort), a NIAID Division of Allergy, Immunology and Transplantation funded data repository under accession code SDY1760 (https://www.immport.org/shared/study/SDY1760). All raw and processed data are available under restricted access to comply with the NIH public data sharing policy for IRB-exempted public health surveillance studies, Access can be obtained via AccessClinicalData@NIAID (https://accessclinicaldata.niaid.nih.gov/study-viewer/clinical_trials). Additional guidelines for access are outlined on ImmPort (https://docs.immport.org/home/impaccslides).

## CODE AVAILABILITY

All codes for the analyses and tables generated by this study have been deposited in the Bitbucket repository https://bitbucket.org/kleinstein/impacc-public-code/src/master/predict_model_PASC_manuscript/ and are publicly available as of the date of publication.

## Acknowledgements

This study is being supported by grants R01AI104870, R01AI132774, R01AI135803, R01AI145835, U19AI057229, U19AI062629, U19AI077439, U19AI089992,U19AI090023, U19AI 118608, U19AI118610, U19AI125357, U19AI128910, U19AI128913, U54AI142766, U19AI089992, U24AI52179 from the National Institute of Allergy and Infectious Diseases (NIAID), a part of the U.S. National Institutes of Health(NIH), and P51 OD011132, S10 OD026799 from NIH.

## IMPACC Network Competing Interests

The Icahn School of Medicine at Mount Sinai has filed patent applications relating to SARS-CoV-2 serological assays and NDV-based SARS-CoV-2 vaccines which list Florian Krammer as co-inventor. Mount Sinai has spun out a company, Kantaro, to market serological tests for SARS-CoV-2. Florian Krammer has consulted for Merck and Pfizer (before 2020), and is currently consulting for Pfizer, Seqirus, 3rd Rock Ventures, Merck and Avimex. The Krammer laboratory is also collaborating with Pfizer on animal models of SARS-CoV-2. Viviana Simon is a co-inventor on a patent filed relating to SARS-CoV-2 serological assays (the “Serology Assays”). Ofer Levy is a named inventor on patents held by Boston Children’s Hospital relating to vaccine adjuvants and human in vitro platforms that model vaccine action. His laboratory has received research support from GlaxoSmithKline (GSK). Charles Cairns serves as a consultant to bioMerieux and is funded for a grant from Bill & Melinda Gates Foundation. James A Overton is a consultant at Knocean Inc. Jessica Lasky-Su serves as a scientific advisor of Precion Inc. Scott R. Hutton, Greg Michelloti and Kari Wong are employees of Metabolon Inc. Vicki Seyfer-Margolis is a current employee of MyOwnMed. Nadine Rouphael reports contracts with Lilly,Immorna, Vaccine Company and Sanofi for COVID-19 clinical trials and serves as a consultant for ICON, EMMES, Imunon, CyanVac for consulting on safety for COVID19 clinical trials. Adeeb Rahman is a current employee of Immunai Inc. Steven Kleinstein is a consultant related to ImmPort data repository for Peraton. Nathan Grabaugh is a consultant for Tempus Labs and the National Basketball Association. Akiko Iwasaki is a consultant for 4BIO, Blue Willow Biologics, Revelar Biotherapeutics, RIGImmune, Xanadu Bio, Paratus Sciences. Monika Kraft receives research funds paid to her institution from NIH, ALA; Sanofi, Astra-Zeneca for work in asthma, serves as a consultant for Astra-Zeneca, Sanofi, Chiesi, GSK for severe asthma; is a co-founder and CMO for RaeSedo, Inc, a company created to develop peptidomimetics for treatment of inflammatory lung disease. Esther Melamed received research funding from Babson Diagnostics, honorarium from Multiple Sclerosis Association of America and has served on advisory boards of Genentech, Horizon, Teva and Viela Bio. Carolyn Calfee receives research funding from NIH, FDA, DOD, Roche-Genentech and Quantum Leap Healthcare Collaborative as well as consulting services for Janssen, Vasomune, Gen1e Life Sciences, NGMBio, and Cellenkos. Wade Schulz was an investigator for a research agreement, through Yale University, from the Shenzhen Center for Health Information for work to advance intelligent disease prevention and health promotion; collaborates with the National Center for Cardiovascular Diseases in Beijing; is a technical consultant to Hugo Health, a personal health information platform; cofounder of Refactor Health, an AI-augmented data management platform for health care; and has received grants from Merck and Regeneron Pharmaceutical for research related to COVID-19. Grace A McComsey received research grants from Rehdhill, Cognivue, Pfizer, and Genentech, and served as a research consultant for Gilead, Merck, Viiv/GSK, and Jenssen. Linda N. Geng received research funding paid to her institution from Pfizer, Inc.

